# Percutaneous aspiration for shortening time to blood cultures sterilization in right-sided infective endocarditis and vegetations

**DOI:** 10.1101/2024.06.12.24308736

**Authors:** Francisco Tirado Polo, Asaad Nakhle, Eric Gnall, Zach Rozenbaum

**Affiliations:** Department of cardiology, Tulane University, New Orleans, Louisiana, USA; Department of cardiology, Lankenau Medical Center, Wynnewood, Pennsylvania, USA

**Keywords:** Percutaneous aspiration, Bacteremia, Infective endocarditis, Vegetations

## Abstract

**Background:** Surgical intervention is associated with earlier clearance of bacteremia in infective endocarditis (IE).

**Aim:** We hypothesized that vegetectomies using percutaneous aspiration shortens time to sterilization of blood cultures in patients with right-sided IE and vegetations.

**Results:** The cohort included 37 patients, 23 treated conservatively, and 14 underwent percutaneous vegetectomy. The median time to blood culture sterilization among patients with bacteremia over 7 days, was 16.5 (IQR 9.75-29) for patients treated conservatively and 11.5 (IQR 8.5-11.5) for those who underwent vegetectomy. The 2 patients who required mechanical ventilation were among the vegetectomy group, and the single patient who died during the same admission was treated conservatively. There were no complications in the vegetectomy group.

**Conclusion:** These data are hypothesis-generating, suggesting that utilizing percutaneous aspiration in patients with right-sided IE and vegetations shortens time to sterilization of blood cultures, and possibly improves outcomes.

## Research letter

Over the past few years, the rate of hospitalization for infective endocarditis (IE) has been increasing(1). Right-side IE occurs predominantly in intravenous drug users (IVDU) and patients with intracardiac devices(2). Although the course of right-sided IE is considered to be more benign than left side IE, in-hospital mortality was found to be 7%(3). Antibiotics are the mainstay of treatment, but mechanical interventions should be utilized in uncontrolled disease, such as persistent bacteremia. Vegetations increase the risk of persistent bacteremia due to the physical barrier that protect bacteria(4). Accordingly, large vegetations are linked to worse outcomes, with up to 33% in-hospital morality(3). Until recent years surgery was the only available mechanical intervention. The possibilities expanded as percutaneous solutions emerged(4). The rationale for using percutaneous aspiration includes avoiding major surgery, allowing sterilization of blood cultures prior to valve replacement/repair if indicated, and in IVUD where reinfection is likely, avoiding implantation of cardiac prosthetic material. Since surgical intervention is associated with earlier clearance of bacteremia(5) we hypothesized that vegetectomies using percutaneous aspiration shortens time to sterilization of blood cultures in patients with right-sided IE and vegetations.

In a single center retrospective study, patients with right-sided IE and vegetations were identified over the course of 3 years 2021-2023. The control group was composed of patients from the first 2 years of the study, when percutaneous aspiration was not available in the hospital. The study group was composed of patients from the last year of the study, when percutaneous aspiration was available. Persistent blood cultures for 7 days is an indication for mechanical intervention(2). Patients were therefore divided into groups according to the time to blood culture sterilization. Additional indications, comprising the group with intervention prior to 7 days, include large vegetations (≥2 cm), resistant pathogens, and recurrent emboli(2). The study was approved by the Tulane University institutional review board with granted waiver of informed consent (Approval number 2023-876-TMC).

The cohort included 37 patients, 23 treated conservatively, and 14 underwent percutaneous vegetectomy. The median age was 40 (IQR 31-48), 43.2% were of female gender, 62.2% had prior IE, and 91.9% were IVDU. The median vegetation size was 1.4 cm (IQR 1-2), 45.9% had ≥severe tricuspid regurgitation, and 13.5% were in shock. The median bacteremia time was 5 days (IQR 3-10), 40.5% had Methicillin-resistant Staphylococcus aureus (MRSA) bacteremia, 10.8% had fungemia, and 70.2% had lung emboli or distant foci.

The median time to blood culture sterilization among patients with bacteremia over 7 days, was 16.5 (IQR 9.75-29) for patients treated conservatively and 11.5 (IQR 8.5-11.5) for those who underwent vegetectomy (Figure 1A). Of those with bacteremia over 7 days, 2 patients had fungemia in the vegetectomy group, and MRSA bacteremia was diagnosed in 3 patients who were treated conservatively and 4 patients who were treated with vegetectomy. The 2 patients who required mechanical ventilation were among the vegetectomy group, and the single patient who died during the same admission was treated conservatively. There were no complications in the vegetectomy group. An example of vegetations removed by percutaneous aspiration is shown in Figure 1B.

**Figure 1:**
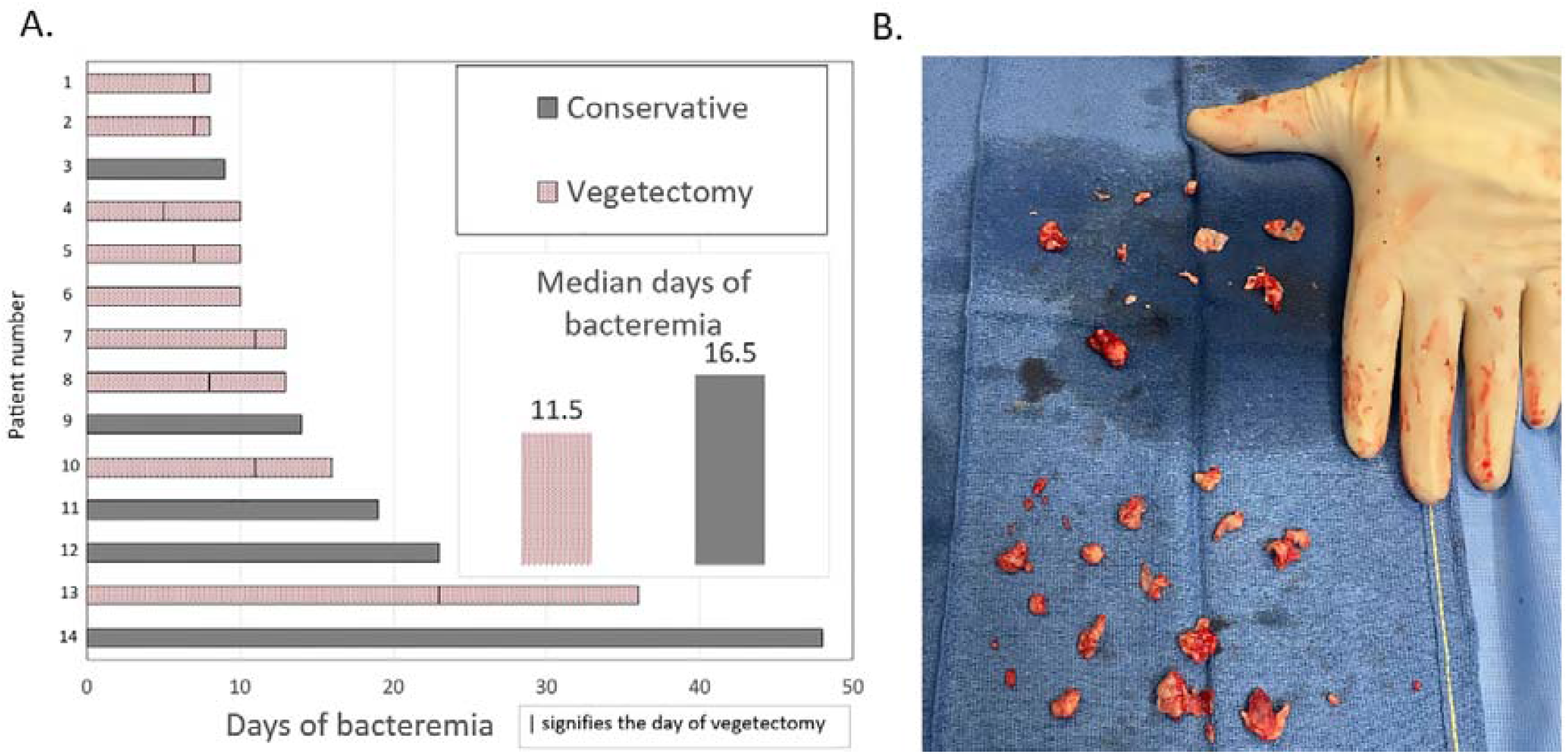
Patients with bacteremia over a week stratified by management strategy A. Days to blood culture sterilization; B. Example of vegetations removed by percutaneous aspiration

This is the first study to compare time to culture sterilization among patients with right-sided IE and vegetations by management strategy. In the current cohort there was a 5-day median decrease in bacteremia among patient treated with percutaneous aspiration. The single patient who died (4%) was treated conservatively, and there was no in-hospital mortality among those who underwent vegetectomy despite more patients with respiratory failure in that group. Of note, 7 patients were discharged against medical advice.

Early intervention in deteriorating patients was previously shown to improve outcomes, however, guidelines recommend avoiding surgery in IVDU(2). Moreover, in unstable patients, surgery may not be feasible. Although large scale data are lacking, the role of percutaneous aspiration is expanding.

The current study is limited by its small sample size. Nevertheless, these data are hypothesis-generating, suggesting that utilizing percutaneous aspiration in patients with right-sided IE and vegetations shortens time to sterilization of blood cultures, and possibly improves outcomes.

## Data Availability

According to the IRB, data may not be shared.

## Notes

### Competing Interest Statement

Dr. Gnall receives advisory fees from Livanova, ABIOMED, and Maquet. Zach Rozenbaum received consultant fees from Angiodynamics. The other co-authors have no disclosures to declare.

### Funding Statement

none

### Author Declarations

The study was approved by the Tulane University institutional review board with granted waiver of informed consent (Approval number 2023-876-TMC).

